# Comparative Analysis of Surrogate Adiposity Markers and Their Relationship With Mortality

**DOI:** 10.1101/2022.09.26.22280347

**Authors:** Irfan Khan, Michael Chong, Ann Le, Pedrum Mohammadi-Shemirani, Robert Morton, Christina Brinza, Michel Kiflen, Sukrit Narula, Loubna Akhabir, Shihong Mao, Katherine Morrison, Marie Pigeyre, Guillaume Paré

**Author notes:** **Address for correspondence:** Guillaume Paré MD, MSc, FRCPC, McMaster University, Population Health Research Institute, David Braley Cardiac, Vascular and Stroke Institute, 237 Barton St. East – C4-126, Hamilton, ON L8L 2X2,Canada.

## Abstract

**Importance:** Body mass index (BMI) is an easily obtainable surrogate for adiposity. However, there is substantial variability in body composition and adipose tissue distribution between individuals with the same BMI. Furthermore, previous literature is conflicting regarding the optimal BMI linked with the lowest mortality risk.

**Objective:** To determine which of BMI, fat mass index (FMI), and waist-to-hip (WHR) is the strongest and most consistent causal predictor for mortality.

**Design:** We created a case-control cohort using all incident deaths from the UK Biobank (UKB; 2006 to 2022).

**Setting:** 22 Clinical assessment centres across the United Kingdom.

**Participants:** We partitioned UKB British participants (N= 387,672) into a discovery (N = 337,078) and validation cohort (N = 50,594), the latter consisting of 25,297 deaths and 25,297 randomly selected age- and sex-matched controls. The discovery cohort was used to derive genetically-determined adiposity measures while the validation cohort was used for all other analyses. Relationships between exposures and outcomes were analyzed through both observational and Mendelian randomization (MR) analyses to infer causality.

**Exposures:** BMI, FMI and WHR.

**Main Outcomes and Measures:** All-cause mortality; Cause-specific mortality (cancer, cardiovascular disease (CVD), respiratory disease, or other causes).

**Results:** Observational relationships between measured BMI and FMI with all-cause mortality were J-shaped, whereas the relationship with WHR was linear. Genetically-determined WHR had a stronger association with all-cause mortality compared to BMI or FMI (OR per SD increase of WHR (95% CI): 1.51 (1.32 – 1.72); 1.29 (1.20 – 1.38) for BMI, and 1.45 (1.36 – 1.54) for FMI, heterogeneity *P*<0.05), and exhibited a stronger effect in males than females (males: 1.89 [1.54 – 2.52], females: 1.20 [1.06-1.30], heterogeneity *P*<0.05). Unlike BMI or FMI, the relationship between genetically-determined WHR and all-cause mortality was consistent irrespective of observed BMI (heterogeneity *P* > 0.05).

**Conclusions and Relevance:** WHR has the strongest and most consistent causal association with risk of mortality irrespective of BMI, with the effect being stronger in males than females. Clinical recommendations and interventions should prioritize adiposity distribution rather than mass.

**Key Points:** *Question:* Among body mass index (BMI), fat mass index (FMI), or waist-to-hip (WHR) ratio, what is the optimal adiposity measure for predicting mortality outcomes in adults?

*Findings:* In this Mendelian randomization study consisting of 387,672 British adult participants from the UK Biobank (UKB), WHR was found to have the strongest and most consistent causal relationship with all-cause and cause-specific mortality.

*Meaning:* WHR was the most robust predictor of mortality risk and may serve as a more appropriate target for health care intervention.

## Introduction

Since the 1980s, the global prevalence of individuals with overweight or obese has doubled to nearly one-third of the world’s population^1^. Current clinical recommendations for treating obesity are largely based on body mass index (BMI). The World Health Organization (WHO) defines a healthy BMI between 18.5 and 24.5 kg/m^2^ based on observational studies that found individuals had the lowest risk of disease or mortality in this range^2,3,4^. However, in recent studies, the BMI range associated with the lowest risk of mortality varies depending on secular trends, ethnicity, and population^5,6,7,8^. Thus, BMI may not be the best measure of health-related adiposity, as higher BMI could either be beneficial or deleterious depending on clinical context. Other measures of adiposity that account for body composition and body fat distribution, such as fat mass index (FMI) or waist-to-hip ratio (WHR) respectively, have been shown to be either strongly associated with BMI or superior to BMI in predicting risk of disease or mortality^9,10,11,12^. It is thus unclear whether BMI is the best clinical measure of adiposity in predicting disease or mortality^12^.

Much of the literature regarding relationships between adiposity and mortality is observational in design. As such, these studies are subject to potential residual confounding and reverse causality biases, creating limitations in inferring causality^9^. Mendelian randomization (MR) uses genetic variants to infer causal relationships between a given exposure (e.g. BMI) and an outcome (e.g. all-cause mortality). MR can accomplish this because genetic alleles are randomized at conception analogous to how participants are shuffled into groups in a randomized controlled trial^9,13^. Indeed, the J-shaped relationship between BMI and all-cause mortality has been reproduced using MR, though it is unclear how body composition and body fat distribution contribute to the J-shaped curve^9,10,12,14^

An optimal adiposity measure for disease risk stratification would have a strong causal relationship with adverse outcomes, consistency across a range of body composition and be easy-to-measure. Additionally, it would accurately quantify the adiposity content associated with the lowest mortality risk. Hence, we perform a comparative analysis between three anthropometric measures of adiposity (BMI, FMI, and WHR) with mortality from all-cause and specific causes, using observational and MR analyses.

### Research Design and Methods

#### Study Population

The UK Biobank (UKB) is a prospective cohort of >500,000 individuals between the ages of 40-69 years^4^. The UKB dataset issued on August 3^rd^, 2021 was used for analyses, including 408,160 unrelated British individuals with genotypic data suitable for analyses. Of these, 20,065 participants with incomplete phenotypic data (e.g. BMI, age, sex) and 423 participants with extreme BMI^3,9,15^(<15 or >50 kg/m^2^) were excluded, arriving at a final sample size of 387,672. We partitioned the British UK Biobank (UKB) population (N= 387,672) into two subsets: a discovery cohort (N=337,078) and a validation cohort (N=50,594) in preparation for deriving genetically-determined adiposity measures through genome-wide association studies (GWAS) and polygenic risk score (PRS) computations. The validation cohort consisted of all-cause mortality cases matched to living controls based on age, sex, and the first 10 principal components (genetic ancestry) according to propensity score, which equilibrates the covariate distribution between cases and controls (N_cases_ = 25,297, N_matched controls_ = 25,297) while the discovery set consisted of the remaining living UKB participants. The partition was done to achieve maximal power. Exposures included BMI, FMI, and WHR, with BMI derived from anthropometric weight and height, FMI measured from bioelectrical impedance, and WHR computed from waist and hip circumference, described previously^16^. FMI was used instead of whole-body fat mass to account for the influence of height in an absolute indicator of adiposity^2^. The outcomes of interest were all-cause, cancer-related, CVD-related, and respiratory disease-related mortality, along with mortality resulting from all other diseases (‘other mortality category’). Outcomes were defined based on the International Classification of Diseases, 10^th^ revision (ICD-10) codes (Supplementary S5)^9^. Prevalent cases of CVD, cancer, and respiratory disease were excluded to avoid reverse causation in all analyses (ICD-10 codes for each disease category is listed in Supplementary S5)^3^.

#### Polygenic risk score calculations

Polygenic risk scores (PRS) are quantitative measures of an individual’s genetic disposition to a certain trait, which are derived from the weighted effects of genetic variants (e.g., single polynucleotide polymorphisms [SNP]) on the phenotype^17^. Individuals with higher scores have higher genetic predisposition to each respective phenotype. GWAS summary statistics for BMI and WHR were obtained from the Genetic Investigation of ANthropometric Traits (GIANT) while summary statistics for FMI were derived from the UKB^4,18,19^. GIANT summary statistics were derived from the Locke et al (2015) meta-analysis, which excluded UKB participants^4^. When deriving summary statistics within the UKB, the GWAS software REGENIE was used on our discovery cohort: briefly, REGENIE uses a linear mixed model to assess the association between genetic variants and a given trait, after adjusting for age, age^2^, chip type, the first 40 genetic principal components (measure for genetic ancestry), and UKB assessment centre, as described elsewhere^20^. To avoid overfitting, when deriving GWAS data for FMI, all UKB participants were split into discovery and validation cohorts as mentioned above, with the GWAS performed in the discovery set only.

Significantly associated genetic variants (*p*-value < 0.01) from each GWAS were used to derive PRS for each exposure (BMI, WHR, and FMI) within each individual (Supplementary S2 and S3)^18,19^. SNPs were pruned for linkage disequilibrium (LD) at a threshold of r^2^ < 0.1^21^. To compute all PRS in the validation dataset (n=50,594), the genetic effect estimates derived from the discovery UKB dataset were applied to individual genotypes using LASSOSUM, a penalized regression method to generate PRS^19,22^. All PRS were standardized for their effects on their corresponding traits (e.g. the BMI PRS was adjusted for its effect on BMI).

#### Observational Analyses

Cox proportional hazard models were used to determine the association between measures of adiposity (BMI, FMI, and WHR) and incident mortality outcomes. Analyses were adjusted for age, sex, and the first 10 genetic principal components. Sex-stratified analyses with all-cause mortality were additionally conducted. ANOVA was used to determine whether a non-linear model provided a better fit than a linear model. If the F-statistic is >10, the non-linear model would have the better fit.All analyses were completed in the full UKB population (N = 387,672).

#### Linear Mendelian Randomization

Linear MR was used to determine the strength of the causal relationship between measures of adiposity (BMI, FMI, and WHR) versus all-cause and cause-specific mortality. Estimates for MR analyses were computed using an allelic scoring method and the Wald ratio. The latter was used to standardize PRS per SD change in adiposity. Three covariate models were used: unadjusted model, which included no covariates; adjusted model including age, sex, and the first 10 genetic principal components as covariates; and adjusted plus other adiposity markers model, where each PRS examined in each analysis was adjusted for the other PRS (e.g., the BMI PRS was controlled for FMI and WHR PRS if analyzing BMI PRS). As secondary analyses, the same models were stratified by sex and presumed menopausal status. Consistent with previous investigations, females aged 52 and younger were defined to be at presumed pre-menopausal age, while presumed post-menopausal age refers to females aged 53 and older^23^.

#### Non-Linear Mendelian Randomization

Non-linear MR (NLMR) was used to determine if the relationships between genetically-determined adiposity (BMI, FMI, and WHR) and mortality outcomes varied depending on an individual’s measured level of adiposity^9,15^. UKB participants were partitioned into 20 quantiles of measured BMI, FMI, or WHR after residualization for the corresponding PRS to avoid collider bias^9,24^. Within each quantile, the Wald ratio for each respective PRS was computed. Ordinary least squares regression was used to determine whether there is variation in the Wald ratio between quantiles, testing for both linearity (i.e. standard linear regression) and non-linearity (i.e. inclusion of a quadratic term). ANOVA was used to test non-linearity for statistical significance. NLMR analyses were performed using the same covariate schemes as previously described for linear MR. All statistical analyses were conducted using R (version 3.6.3). Statistical significance was set at two-tailed *p* values < 0.05.

## Results

### Observational associations between adiposity and mortality

The baseline characteristics of study participants are shown in Supplementary S1. A J-shaped association was found between both measured BMI and FMI with all-cause mortality, with the nadir at a BMI of 24.9 kg/m^2^ and an FMI of 6.15 kg/m^2^ (*P*_non-linearity_ = 4.71 × 10^−169^ and 1.90 × 10^−33^ ; Figure 1). A significant monotonically increasing relationship was found between WHR and all-cause mortality (Figure 1). These relationships were consistent when analyzing males and females separately (Supplementary S10).

**Figure 1.**
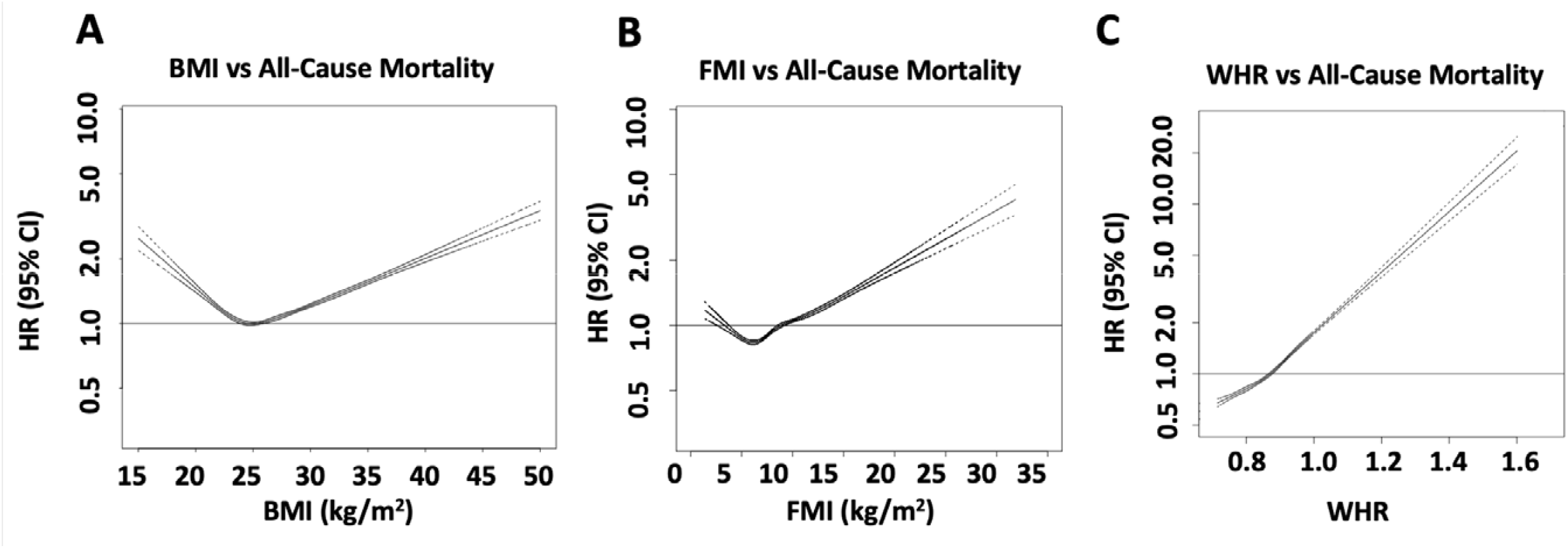
A) BMI-, B) FMI-, and C) WHR- all-cause mortality relationships in all participants (N= 387,672). BMI = body mass index, FMI = fat mass index, WHR = Waist-to-hip ratio, HR = hazard ratio for all-cause mortality. Statistical significance for non-linearity is *p* < 0.05. The reference point at HR = 1 for BMI (25 kg/m^2^), the mean value for FMI in the UKB population (8.83 kg/m^2^), and the mean value for WHR in the UKB population (0.87) for analyses with BMI, FMI, and WHR were used as independent variables, respectively. The nadir for BMI and FMI was 24.9 and 6.15 kg/m^2^, respectively. The WHR associated with an HR of 1 was 0.87.

Observational analyses show that there were positive relationships for the three adiposity measures with cancer mortality (OR per SD change in adiposity [95% CI]: BMI: 1.06 [1.04 – 1.08], *p* < 0.05; FMI: 1.08 [1.05 – 1.10], *p* < 0.05; WHR: 1.18 [1.15 – 1.21], *p* < 0.05) and CVD mortality (OR per SD change in adiposity [95% CI]: BMI: 1.41 [1.37 – 1.45], *p* < 0.05; FMI: 1.45 [1.40 – 1.49], *p* < 0.05; WHR: 1.59 [1.53 – 1.64], *p* < 0.05). For BMI and FMI, there were J-shaped associations with mortality due to respiratory disease and ‘other’ causes of mortality. For respiratory disease-associated mortality, the nadirs corresponding to the adiposity levels associated with the lowest rate of mortality were 26.0 and 7.43 kg/m^2^, for BMI and FMI, respectively. For the other mortality category, the nadirs for BMI and FMI were 25.5 and 6.55 kg/m^2^, respectively. In contrast, WHR exhibited monotonically increasing associations with cause-specific mortality (Supplementary S11).

### Mendelian randomization associations of adiposity with mortality

To verify the relevance of PRS, we confirmed that all PRS were significantly associated with their corresponding traits in the independent validation set (Supplementary S6). There was a significant causal relationship between all three adiposity measures and all-cause mortality (OR per SD change in genetically-determined adiposity measure [95% CI]: BMI: 1.29 [1.20 – 1.38], *p* = 1.44 × 10^−13^; FMI: 1.45 [1.36 – 1.54], *p* = 6.27 × 10^−30^ ; WHR: 1.51 [1.32 – 1.72], *p* = 2.11 × 10^−9^) (Table 1). WHR had the strongest association with all-cause mortality (*p* < 0.05 for all comparisons, Figure 2). Causal relationships were significant irrespective of adjustment (*p* < 0.05, Table 1).

**Table 1.**
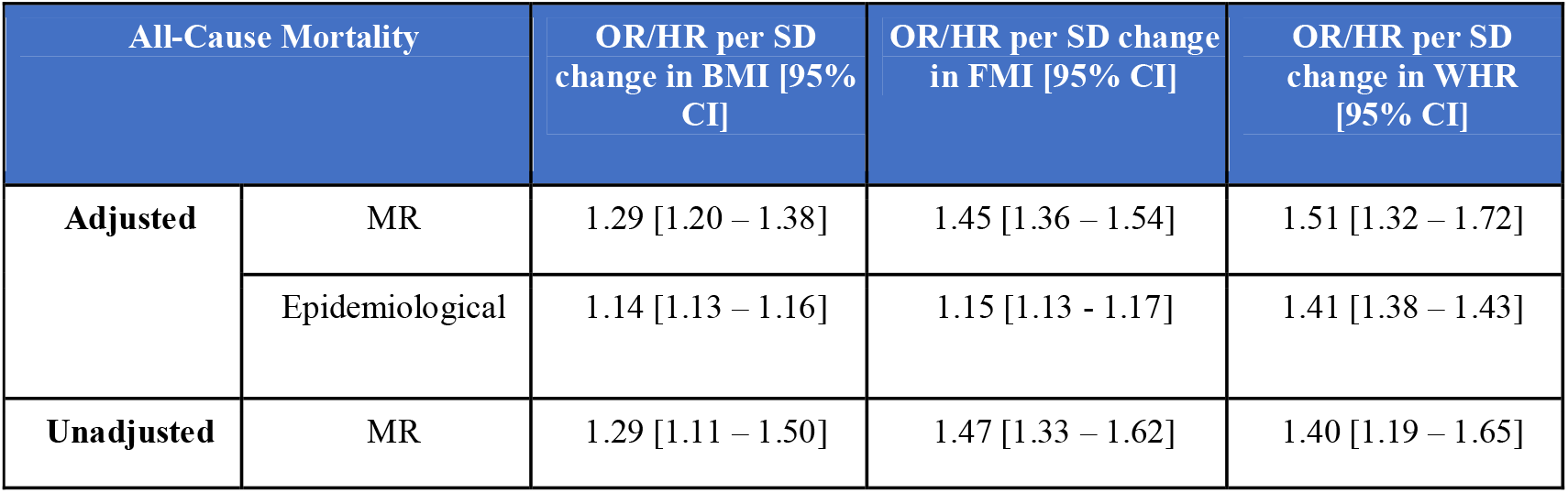
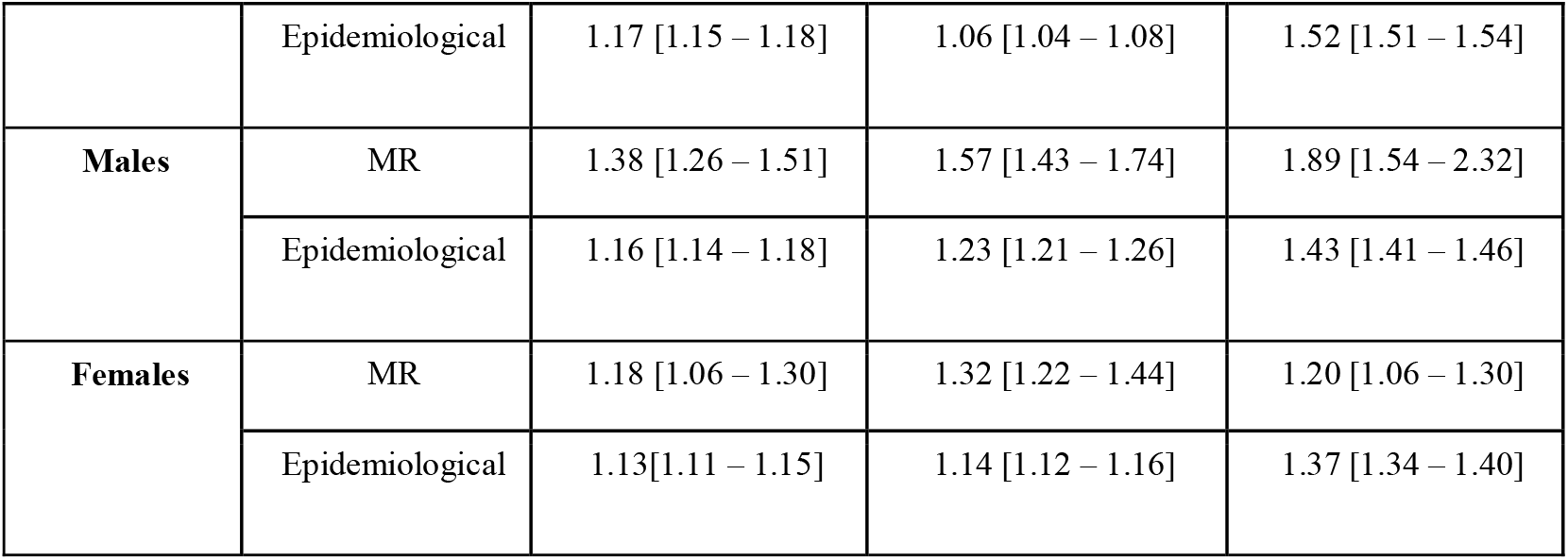
Summary of epidemiological and linear mendelian randomization analyses of adiposity measures and all-cause mortality. Hazard ratios (HR) indicate the effect of a 1 SD unit increase in adiposity measure on risk of all-cause mortality. Odds ratios (OR) indicate the effect of a 1 SD unit increase in genetically-determined adiposity measure on risk of all-cause mortality. The adjusted model was used for all sex-specific analyses. MR = mendelian randomization, BMI = body mass index, FMI = fat mass index, WHR = waist-to-hip ratio, OR = odds ratio, HR = hazard ratio, PRS = polygenic risk score. N=50,594 (adjusted model with males and females combined), N=30,031 (males only cohort), and N=20,563 (females only cohort). All relationships have *p* < 0.05.

**Figure 2.**
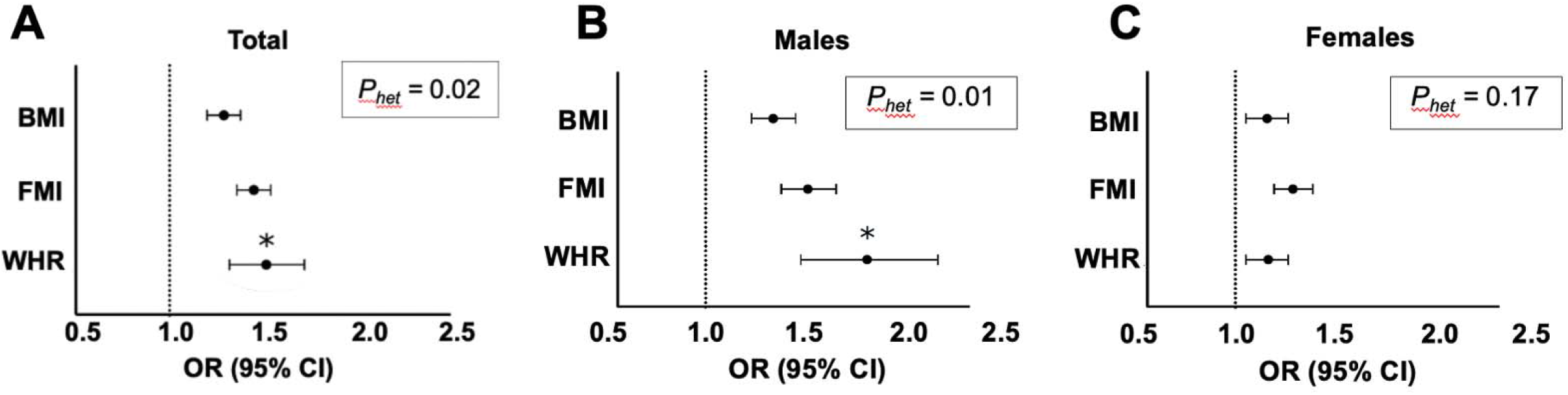
Linear mendelian randomization analyses comparing genetically-determined adiposity measures in their relationship with all-cause mortality. The minimally adjusted model was used in the total UKB population representing all UKB participants, as well as males and females only. All PRS were standardized for their effects on their corresponding traits (e.g. the BMI PRS was adjusted for its effect on BMI). Odds ratios (OR) indicate the effect of a 1 SD unit increase in adiposity measure on risk of all-cause mortality. Significance is considered at *p* < 0.05. BMI = body mass index, FMI = fat mass index, WHR = waist-to-hip ratio, OR = odds ratio, PRS = polygenic risk score, *P*_het_ = *p* value for general heterogeneity from the fixed-effects heterogeneity test. Asterisks (*) represent statistical significance compared to the other adiposity measures. N_total_ = 50,007, N_males_ = 30,031, N_females_ = 20,563.

MR analyses were also performed separately for males and females (Table 1 and Figure 2). There was a significant interaction between sex and genetically-determined adiposity measures with mortality outcomes (Table 1, *p* < 0.05 for all three measures). A significant association between WHR and all-cause mortality was observed in males (OR [95% CI]: 1.89 [1.54 – 2.32], *p* = 1.44 × 10^−9^, Table 1) and females (OR [95% CI]: 1.20 [1.01 – 1.43], *p* = 3.77 × 10^−2^, Table 1). In males, pairwise heterogeneity comparison analyses demonstrated that WHR had a stronger effect on all-cause mortality than either BMI or FMI (heterogeneity *p*<0.05 for both comparisons, Table 1 and Figure 2). In females, the three measures of adiposity were similarly related to all-cause mortality (heterogeneity *p*>0.05 for both comparisons, Table 1 and Figure 2). There was no significant difference between females of pre- or post-menopausal age in the causal effect of the three adiposity measures on all-cause mortality (heterogeneity *p* > 0.05, Supplementary S12).

To test the causal relationship between adiposity measures and cause-specific mortality, MR analyses were repeated among participants with cancer, CVD, respiratory disease, and other disease mortality. All adiposity measures were significantly associated with mortality due to CVD or other diseases (*p* < 0.05 for all, Table 2). Only FMI was significantly associated with respiratory disease mortality (*p* < 0.001, Table 2). There were no significant differences between the three adiposity measures in their effect on the four mortality outcomes (*P*_*het*_ > 0.05, Table 2).

**Table 2.** Summary of epidemiological and linear mendelian randomization analyses of adiposity measures and cause-specific mortality. Hazard ratios (HR) indicate the effect of a 1 SD unit increase in adiposity measure on risk of cause-specific mortality. Odds ratios (OR) indicate the effect of a 1 SD unit increase in genetically-determined adiposity measure on risk of cause-specific mortality. The adjusted model was used for all analyses involving cause-specific mortality. Significance is considered at *p* < 0.05. MR = mendelian randomization, BMI = body mass index, FMI = fat mass index, WHR = waist-to-hip ratio, OR = odds ratio, HR = hazard ratio, PRS = polygenic risk score, CVD = cardiovascular disease. N=50,594.

| Cause-Specific Mortality |  | OR/HR per SD change in BMI [95% CI] | P value | OR/HR per SD change in FMI [95% CI] | P value | OR/HR per SD change in WHR [95% CI] | P value |
| --- | --- | --- | --- | --- | --- | --- | --- |
| **Cancer** | MR | 1.05 [0.97 - 1.15] | 0.23 | 1.01 [0.93 -1.09] | 0.84 | 1.04 [0.87-1.23] | 0.68 |
|  | Epidemiological | 1.06 [1.04 - 1.08] | < 0.001 | 1.07 [1.05 -1.09] | < 0.001 | 1.18 [1.15 - 1.21] | < 0.001 |
| **CVD** | MR | 1.30 [1.15- 1.47] |  | 1.38 [1.23 -1.55] |  | 1.56 [1.23-1.99] |  |
|  | Epidemiological | 1.41 [1.37- 1.45] |  | 1.45 [1.39 -1.51] |  | 1.59 [1.53-1.64] |  |
| **Respiratory** | MR | 1.00 [0.83 - 1.19] | 0.96 | 1.33 [1.12 - 1.59] | < 0.001 | 1.11 [0.77 - 1.60] | 0.57 |
|  | Epidemiological | 0.91 [0.87 - 0.96] | < 0.001 | 0.99 [0.93- 1.05] | < 0.001 | 1.26 [1.18- 1.34] | < 0.001 |
| **Other** | MR | 1.26 [1.17- 1.34] |  | 1.40 [1.31 - 1.49] |  | 1.39 [1.22- 1.60] |  |
|  | Epidemiological | 1.10 [1.07- 1.14] |  | 1.12 [1.07 - 1.16] |  | 1.32 [1.27- 1.37] |  |

Using the adjusted model, there were differences between MR and epidemiological effect estimates for the BMI, FMI, and WHR–mortality relationships, with MR estimates having stronger strengths of association with mortality (*p* < 0.05, Supplementary S12, S13, and S14).

### Non-linear mendelian randomization analyses

To determine whether the causal effect of adiposity was constant across different levels of observed adiposity and body shape, we performed non-linear MR analyses. The effect of genetically-determined BMI and FMI on all-cause mortality varied across quantiles of observed BMI, but WHR did not (*p* = 0.04, *p* = 0.03, and *p =* 0.05, Figure 3 respectively).

**Figure 3.**
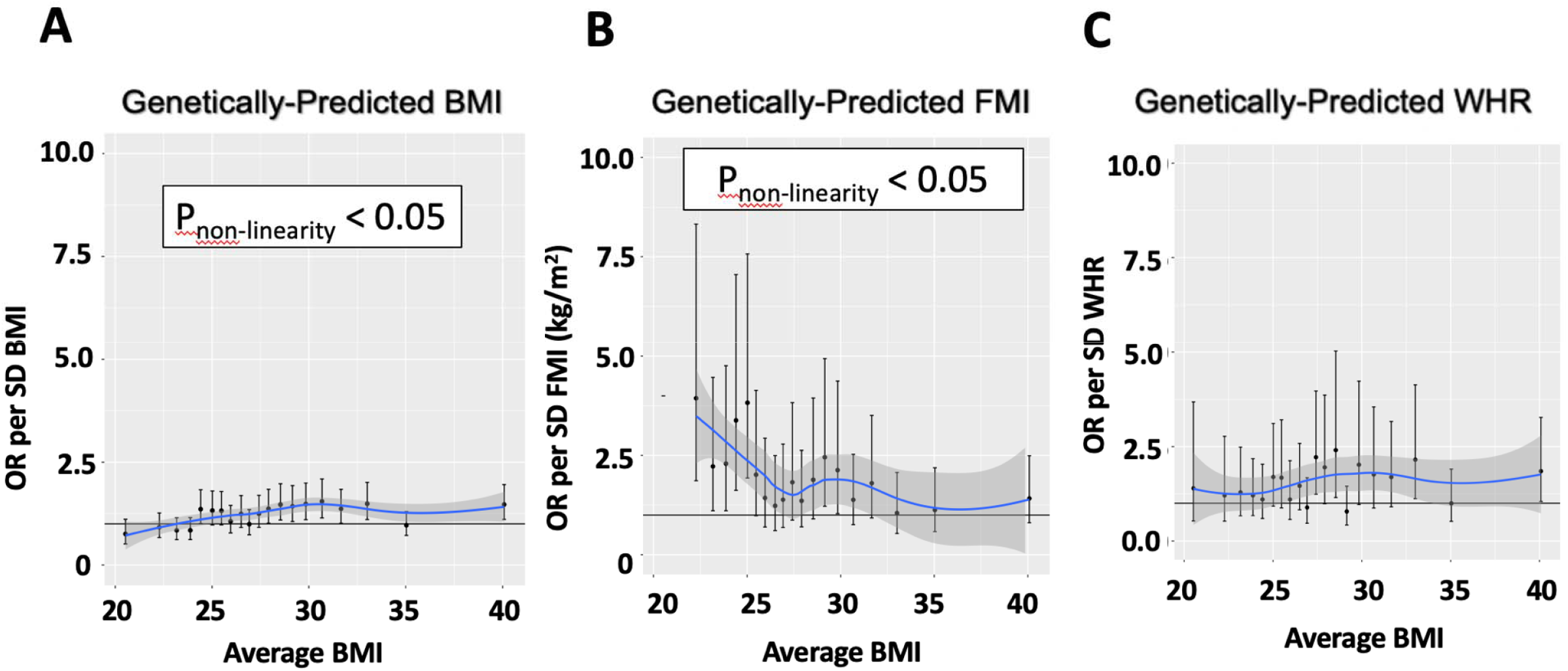
Non-linear mendelian randomization analyses comparing genetically-determined adiposity measure-all-cause mortality relationships across BMI quantiles. All PRS were standardized for their effects on their corresponding traits (i.e. the BMI PRS was adjusted for its effect on BMI). Significance is considered at p < 0.05. Asterisks (*) represent statistical significance. Statistical significance indicates inconsistency of the adiposity-mortality relationship across quantiles of the relevant adiposity measure. The horizontal line indicates an OR of 1. BMI = body mass index, FMI = fat mass index, WHR = waist-to-hip ratio, PRS = polygenic risk score, OR= odds ratio for all-cause mortality per SD of the adiposity measure, *P*_non-linearity_ = *p* value from ANOVA testing for non-linearity. n = 50,007.

## Discussion

An optimal marker of adiposity should be easy to measure and meet the following three characteristics: causal relationships, strong association with mortality, and consistency across adiposity levels. Our study revealed that the relationship between WHR and all-cause mortality is causal, stronger than BMI or FMI and is consistent across quantiles of BMI. While WHR is considered a simple measure, it is as easy to measure as BMI^25^. Both measures are easier to measure than FMI due to FMI requiring BIA, which can be expensive and inaccessible^16,25^.

Our MR findings support a causal relationship between WHR and risk of mortality. This suggests that WHR can not only be used for population health surveillance as a marker of risk, but also that it may be a suitable target for intervention. We also found an association using MR between WHR and mortality from both CVD and other diseases, which is broadly consistent with a prior study that found genetically-determined WHR was associated with higher levels of 2-hour glucose, triglycerides, and systolic blood pressure as well as risk of type 2 diabetes (T2D) and coronary heart disease (CHD)^10^. These findings provide a potential causal pathway linking WHR to mortality and especially cardiovascular mortality^9^. Nevertheless, having a causal relationship with mortality is not sufficient for an adiposity measure to be considered optimal. Indeed, causal relationships for BMI and FMI with all-cause mortality were also supported by MR analyses.

Both our epidemiological and MR analyses also demonstrated that WHR was a stronger predictor of all-cause mortality than FMI or BMI. This result is consistent with previous literature where BMI, although widely used in clinical practice, has been shown to be weaker at predicting disease or mortality risk compared to FMI or WHR^9,10,11,12^. In one MR study examining the causal effect of genetically-determined BMI and WHR on blood pressure traits, WHR was found to have a larger effect than BMI, despite both measures being significantly associated with these traits^26^.

An optimal marker of adiposity should also provide consistent associations across different adiposity levels. Contrary to BMI and FMI, the association of WHR with mortality did not differ across BMI quantiles. When contextualizing this result to the lower end of the J-shaped BMI- mortality curve, our results show that increased abdominal fat mass is still detrimental to health, despite low BMI. Indeed, previous studies have postulated that the lower end of the J-shape curve could be driven by the loss of lean mass due to disease-related cachexia or malnutrition, regardless of the amount of FM and/or abdominal adiposity still present^27^. This makes WHR the superior adiposity measure: it is not only robust to variation in BMI, but also simplifies the clinical recommendations in that lower WHR universally leads to better health outcomes.

Our study showed that males with a higher WHR have a disproportionately higher risk of mortality than females. Thus, assessing and targeting WHR may have greater relevance in males than females. Since female adiposity distribution is drastically different to males, the relatively higher abdominal obesity in males may explain the sex-specific difference in mortality risk^2,28,29^. A previous MR study showed that increased WHR was linked with larger effects on risk of renal outcomes, a leading cause of death, in males compared to females^29^. We did not observe a difference between females of pre- and post-menopausal age, although we were limited by a relatively small sample size in the pre-menopausal aged group compared to the post-menopausal age group. Future studies would be required to reliably assess a menopause-age related interaction.

A few limitations should be mentioned. We used a genetically homogeneous and unrelated British Caucasian population, such that our results may not be representative of other populations. Future studies in larger non-European populations should be conducted to see whether our findings persist in other ethnic groups^6^. All adiposity measures were assessed at baseline and we could not assess whether changes in adiposity over time impact mortality. However, while the UKB does have follow-up data on some adiposity measures, the sample size is relatively small and thus not amenable to such an investigation. Bioelectrical impedance (BIA) was used to measure FMI as opposed to the gold standard dual-energy X-ray absorptiometry (DEXA)^30^. BIA may have limited measurement accuracy and thus, strength of the corresponding mortality associations compared to DEXA^30^.

However, DEXA is expensive and more complicated to use in the clinical setting compared to WHR^30^. Thus, even if we used DEXA for FMI, WHR would still be superior, as it is stronger, simpler, and less expensive to use clinically. Strengths of our study include a large homogenous sample size and an innovative and robust non-linear MR method to establish consistency across the body composition.

Compared to BMI and FMI, WHR had the strongest association with all-cause mortality and was the only measurement that was unaffected by BMI. Indeed, the WHR relationship with all-cause mortality was nearly twice the magnitude of the BMI relationship. Our study provides strong evidence that WHR is a more robust and consistent prognosticator of mortality than BMI. Current WHO recommendations for optimal BMI range are inaccurate across individuals with various body compositions and therefore suboptimal for clinical appraisal and guidelines. Future research is needed to explore whether using WHR as the primary clinical measure of adiposity help improve long-term health outcomes in distinct patient populations as compared to BMI. Independent of BMI, our results provide further support to shift public health focus from measures of general adiposity such as BMI to adiposity distribution using WHR.

## Data Availability

Individual-level UKBiobank genotypes and phenotypes can be acquired upon successful application (https://bbams.ndph.ox.ac.uk/ams/). All individual-level UKBiobank data was accessed as part of application # 15255. GIANT summary statistics are freely available to download (https://portals.broadinstitute.org/collaboration/giant/index.php/GIANT_consortium). All data products generated as part of this study will be made publicly accessible. All remaining data are available in the main text or supplementary materials.

