## Supplementary Material for "Comparative Analysis of Surrogate Adiposity Markers and Their Relationship With Mortality"

**Supplementary for Waist-to-hip ratio is a stronger, more consistent predictor of all-cause mortality than BMI**

**Table of Contents**

Supplementary S1: Baseline characteristics of participants in the UK Biobank (UKB)

Supplementary S2: List of GWAS consortia

Supplementary S3: UKB population and extended methods

Supplementary S4: Body Mass Index (BMI) association with covariates

Supplementary S5: Phenotype and outcome definitions

Supplementary S6: Relationship of BMI, fat mass index (FMI), and waist-to-hip ratio (WHR) with BMI, FMI, and WHR PRS

Supplementary S7: Collinearity between BMI, FMI, and WHR traits

Supplementary S8: Collinearity between BMI, FMI, and WHR PRS

Supplementary S9: Validation of BMI, FMI, and WHR PRS

Supplementary S10: The epidemiological relationship between BMI, FMI, and WHR with all-cause mortality in all UKB participants males, and females

Supplementary S11: The epidemiological relationship between BMI, FMI, and WHR respectively, and cause-specific mortality outcomes

Supplementary S12: Linear mendelian randomization analyses comparing the effect of individual genetically-determined adiposity measures on all-cause mortality between pre versus post-menopausal aged females in the UKB.

Supplementary S13: Comparison between epidemiologically-derived and MR-derived estimates for the BMI – all-cause mortality relationship.

Supplementary S14: Comparison between epidemiologically-derived and MR-derived estimates for the FMI – all-cause mortality relationship.

Supplementary S15: Comparison between epidemiologically-derived and MR-derived estimates for the WHR – all-cause mortality relationship.

**Supplementary S1: Baseline characteristics of participants in the UKB**

Whole UKB Population

| Baseline characteristics | All UKB participants | All deaths | Controls* |
| --- | --- | --- | --- |
| Number of participants | 387,672 | 29,058 | 337,666 |
| Percentage (%) of men | 177,346 (45.7) | 13,732 (59.5) | 163,614 (44.9) |
| Mean (SD) age at baseline (years) | 56.9 (8.0) | 61.8 (6.3) | 56.6 (8.0) |
| Mean (SD) body mass index (kg/m^2^) | 27.4 (4.6) | 28.1 (5.1) | 27.3 (4.6) |
| Mean (SD) whole body fat mass (kg) | 24.8 (9.3) | 25.6 (10.0) | 24.8 (9.3) |
| Mean (SD) height (m) | 1.69 (9.23) | 1.69 (9.22) | 1.69 (9.23) |
| Mean (SD) fat mass index (kg/m^2^) | 8.83 (3.55) | 9.06 (3.74) | 8.82 (3.55) |
| Mean (SD) waist-to-hip ratio | 0.87 (0.1) | 0.91 (0.1) | 0.87 (0.1) |
| Death due to cancer | N/A | 9,732 | 377,940 |
| Death due to cardiovascular disease (CVD) | N/A | 4,231 | 383,441 |
| Death due to respiratory disease | N/A | 1,755 | 385,917 |
| Death due to non-cancer, CVD, or respiratory disease (“other”) | N/A | 4,653 | 383,019 |

*The pure control group, which excludes all mortality cases and controls matched to mortality cases.

Case-Control Sample (i.e. The Testing or Validation Set)

| Baseline characteristics | All UKB participants | All deaths | Controls* |
| --- | --- | --- | --- |
| Number of participants | 50,594 | 25,297 | 25,297 |
| Percentage (%) of men | 30,031 (59.3) | 15,016 (59.3) | 15,015 (59.4) |
| Mean (SD) age at baseline (years) | 61.6 (6.2) | 61.8 (6.2) | 61.3 (6.1) |
| Mean (SD) body mass index (kg/m^2^) | 27.9 (4.8) | 28.2 (5.1) | 27.6 (4.4) |
| Mean (SD) whole body fat mass (kg) | 25.1 (9.5) | 25.7 (10.0) | 24.5 (8.9) |
| Mean (SD) height (m) | 1.70 (9.19) | 1.69 (9.22) | 1.70 (9.16) |
| Mean (SD) fat mass index (kg/m^2^) | 8.83 (3.57) | 9.06 (3.74) | 8.61 (3.37) |
| Mean (SD) waist-to-hip ratio | 0.90 (0.1) | 0.91 (0.1) | 0.89 (0.1) |
| Death due to cancer | N/A | 9,732 | 40,862 |
| Death due to cardiovascular disease (CVD) | N/A | 4,231 | 46,363 |
| Death due to respiratory disease | N/A | 1,755 | 48,839 |
| Death due to non-cancer, CVD, or respiratory disease (“other”) | N/A | 4,653 | 45,941 |

**Supplementary S2: List of GWAS Consortia**

| **Phenotype** | **Consortia Name** |
| --- | --- |
| BMI | Genetic Investigation of ANthropometric Traits (GIANT)^1^ |
| Waist-to-hip ratio (WHR) | Genetic Investigation of ANthropometric Traits (GIANT)^1^ |

**Supplementary S3: UKB population and extended methods**

*UKB Population*

We used the UKB dataset issued on August 3rd, 2021 as part of approved application #15255. Samples were excluded based on criteria from the standard UKB genomic analysis exclusion list (i.e. UKB data showcase data-field #22010)^13^. Further quality control (QC) was also done: related samples, samples with discordant reported sex versus genetic sex, lacking British ancestry, and with QC failure in the UK BiLEVE array were removed^13^. SNPs that had low minor allele frequency (MAF < 0.01), low quality of imputation (INFOscore < 0.6), deviation from Hardy-Weinberg equilibrium/poor genotype calling (HWE; *P* < 10^-6^) or low call rates (< 99%) were removed^13^.

**Schematic representation of sample eligibility criteria and partitioning:**

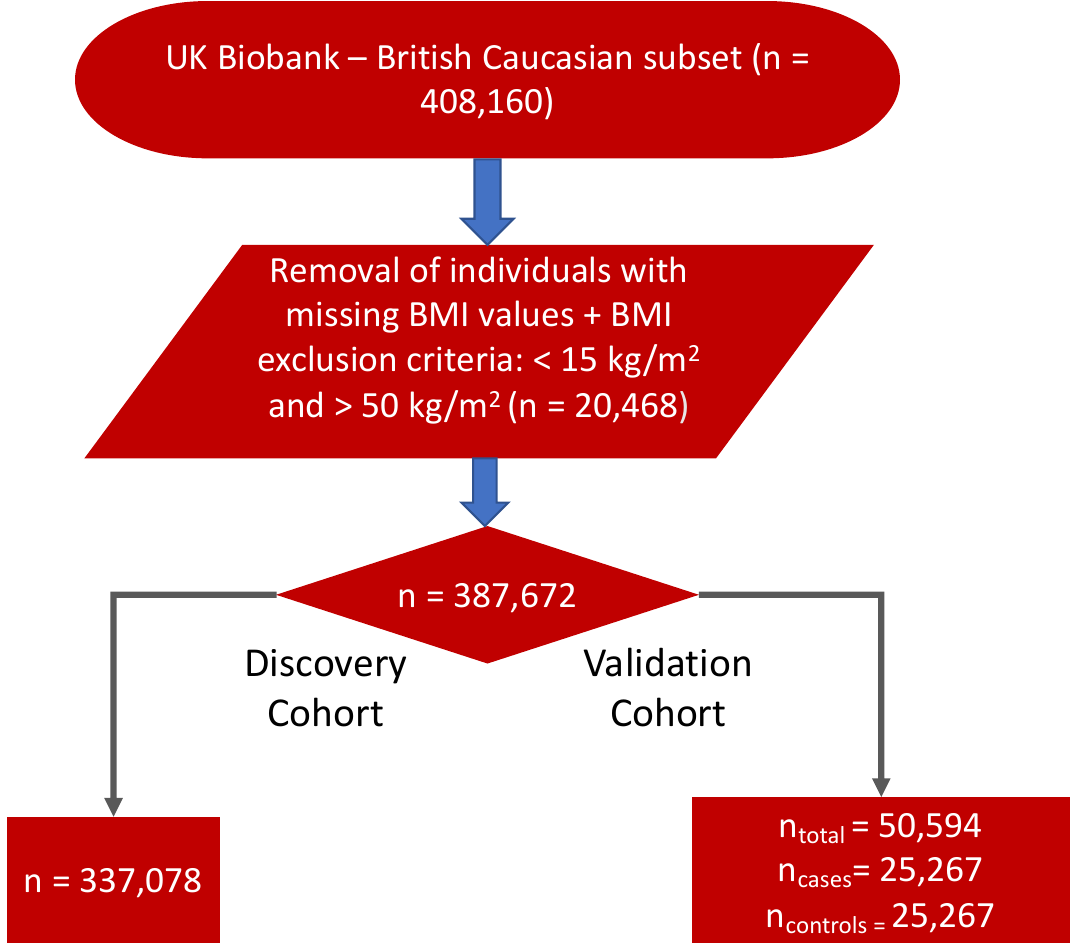

*UK Biobank-Based Genome-Wide Association Studies*

REGENIE, a program that uses whole genome regression modelling to run GWAS analyses, was used to compute GWAS within a select subset of UKB participants for fat mass index (FMI or whole body fat mass/height^2^). REGENIE uses linear regression to assess the association between genetic variants and a given trait, after adjusting for age, age^2^, chip type, the first 40 genetic principal components, and UKB assessment centre, as described elsewhere^9,19^.

*Polygenic risk score calculations*

Polygenic risk score (PRS) was calculated based on the following equation: $\sum_{i=1}^{d} x_{i,d} \frac{b_{d}}{\eta_{d}}$, where $x_{i,d}$ represents the normalized genotype (i.e. the number of risk alleles) per single nucleotide polymorphism (SNP) *d*, $b_{d}$ represents the beta regression coefficient per SNP *d* (e.g. with the BMI SNPs, expressed as a kg/m^2^ increase per risk allele), and $\eta_{d}$ was the linkage disequilibrium (LD) adjustment per SNP *d*^4^. The LD adjustment is based on the sum of the pairwise disequilibrium per SNP *d* over the surrounding 200 SNPs upstream and downstream^4^. PRS were generated from variants of each metabolic phenotype. LASSOSUM was used to generate PRS: it is a method that uses a non-Bayesian penalized regression, as described elsewhere^16^. Associations of all PRS with their respective phenotypes were performed in our sample using regression models (Supplementary S4, S6-9).

The summary statistics from GWAS consortia data in Supplementary S2 was used to generate the BMI and WHR PRS. The summary statistics derived from the UKB-based GWAS analyses for the FMI trait was used to compute the FMI PRS in the validation cohort consisting of mortality cases and matched controls only.

**Supplementary S4: BMI association with covariates**

| **Covariate ~ BMI Association** | **β/OR** | ***P* value** | **Adjusted R-squared** |
| --- | --- | --- | --- |
| Age ~ BMI | 0.01 | 0.09 | 3.67 x 10^-5^ |
| Sex ~ BMI | 0.02 | 1.03 x 10^-37^ | N/A |
| PC1 ~ BMI | -0.003 | 0.02 | 8.11 x 10^-5^ |
| PC2 ~ BMI | -0.0002 | 0.86 | -1.91 x 10^-5^ |
| PC3 ~ BMI | -0.001 | 0.32 | -5.74 x 10^-7^ |
| PC4 ~ BMI | 0.01 | 0.0005 | 0.0002 |
| PC5 ~ BMI | 0.03 | 5.42 x 10^-5^ | 0.0003 |
| PC6 ~ BMI | -0.003 | 0.05 | 5.82 x 10^-5^ |
| PC7 ~ BMI | 0.0006 | 0.69 | -1.67 x 10^-5^ |
| PC8 ~ BMI | -0.002 | 0.20 | 1.24 x 10^-5^ |
| PC9 ~ BMI | -0.01 | 0.02 | 9.43 x 10^-5^ |
| PC10 ~ BMI | 0.002 | 0.44 | -8.05 x 10^-6^ |

* Significance set at *p* < 0.05

**Estimates derived from the testing/validation cohort.

**Supplementary S5: Phenotype and mortality outcome definitions**

| **Phenotype** | **UKB Field ID/ICD-10 Code** |
| --- | --- |
| BMI | 21001-0.0 |
| Sex | 31-0.0 |
| Age | 21022-0.0 |
| FMI | 23100-0.0 and 50-0.0 |
| WHR | 48-0.0 and 49-0.0 |
| Height | 50-0.0 |

| **Mortality Outcome** | **UKB Field ID/ICD-10 Code** |
| --- | --- |
| All-Cause Mortality | 40000-0.0 |
| Cardiovascular Mortality | ICD-10 Code: I |
| Cancer Mortality | ICD-10 Code: C |
| Respiratory Disease Mortality | ICD-10 Codes: J00-09, J10-19, J20-22, J23-29, J3-9 |

**Supplementary S6: Relationship of BMI, FMI, and WHR with BMI, FMI, and WHR PRS**

All regression analyses were adjusted for PRS other than the one being analyzed (e.g. the BMI ~ BMI PRS analysis was adjusted for FMI PRS and WHR PRS), age, sex, and the first 10 principal components.

| **Phenotype ~ BMI PRS Association** | **β** | ***P* value ***** |
| --- | --- | --- |
| BMI ~ BMI PRS | 0.20 | 0.004 |
| FMI ~ BMI PRS | 0.09 | 0.005 |
| WHR ~ BMI PRS | -0.04 |  |

| **Phenotype ~ FMI PRS Association** | **β** | ***P* value ***** |
| --- | --- | --- |
| BMI ~ FMI PRS | 0.20 | 0.005 |
| FMI ~ FMI PRS | 0.25 |  |
| WHR ~ FMI PRS | 0.08 |  |

| **Phenotype ~ WHR PRS Association** | **β** | ***P* value ***** |
| --- | --- | --- |
| BMI ~ WHR PRS | 0.02 | 0.007 |
| FMI ~ WHR PRS | 0.03 |  |
| WHR ~ WHR PRS | 0.25 |  |

* Significance set at *p* < 0.05

**Estimates derived from the testing/validation cohort.

**Supplementary S7: Collinearity between BMI, FMI, and WHR traits**

| **Phenotype ~ Phenotype Association** | **β** | ***P* value***** | **Adjusted R-squared** |
| --- | --- | --- | --- |
| BMI ~ WHR | 22.9 | < 2 x 10^-16^ | 0.19 |
| BMI ~ FMI | 1.12 | < 2 x 10^-16^ | 0.71 |
| FMI ~ WHR | 3.62 | < 2 x 10^-16^ | 0.008 |

* Significance set at *p* < 0.05

**Estimates derived from the testing/validation cohort.

**Supplementary S8: Collinearity between BMI, FMI, and WHR PRS**

| **PRS ~ PRS Association** | **β** | ***P* value ***** | **Adjusted R-squared** |
| --- | --- | --- | --- |
| BMI PRS ~ WHR PRS | 0.48 | < 2 x 10^-16^ | 0.23 |
| BMI PRS ~ FMI PRS | 0.80 | < 2 x 10^-16^ | 0.65 |
| FMI PRS ~ WHR PRS | 0.41 | < 2 x 10^-16^ | 0.17 |

* Significance set at *p* < 0.05

**Estimates derived from the testing/validation cohort.

**Supplementary S9: Validation of BMI, FMI, and WHR PRS**

The following regression models were not adjusted for PRS other than the one being analyzed, age, sex, and the first 10 principal components, unlike Supplementary S6.

| **Phenotype ~ PRS Association** | **β** | ***P* value***** | **Adjusted R-squared** |
| --- | --- | --- | --- |
| BMI ~ BMI PRS | 1.24 | < 2 x 10^-16^ | 0.07 |
| FMI ~ FMI PRS | 1.27 | < 2 x 10^-16^ | 0.11 |
| WHR ~ WHR PRS | 0.01 | < 2 x 10^-16^ | 0.02 |

*Significance set at *p* < 0.05

**Estimates derived from the testing/validation cohort.

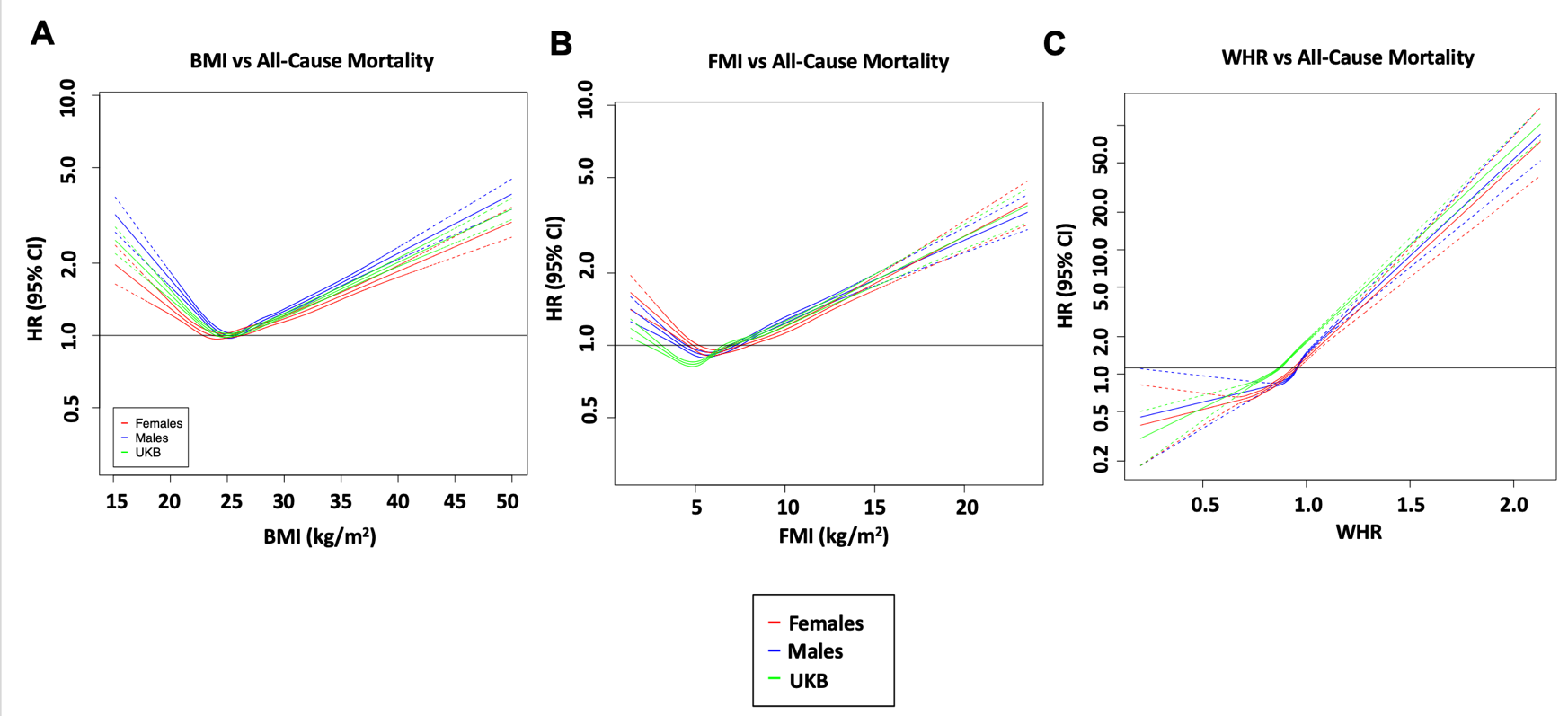

**Supplementary S10:** The relationship between a) BMI, b) FMI, and c) WHR with all-cause mortality in all UKB participants (N=387,672), males (N=177,340), and females (N=210,332). BMI = body mass index, FMI = fat mass index, WHR = Waist-to-hip ratio, HR = hazard ratio for all-cause mortality. Statistical significance for non-linearity Bonferroni-corrected to *p* < 0.05. The reference point at HR = 1 for BMI (25 kg/m^2^), the mean value for FMI in the UKB population (8.83 kg/m^2^), and the mean value for WHR in the UKB population (0.87) for analyses with BMI, FMI, and WHR were used as independent variables, respectively.

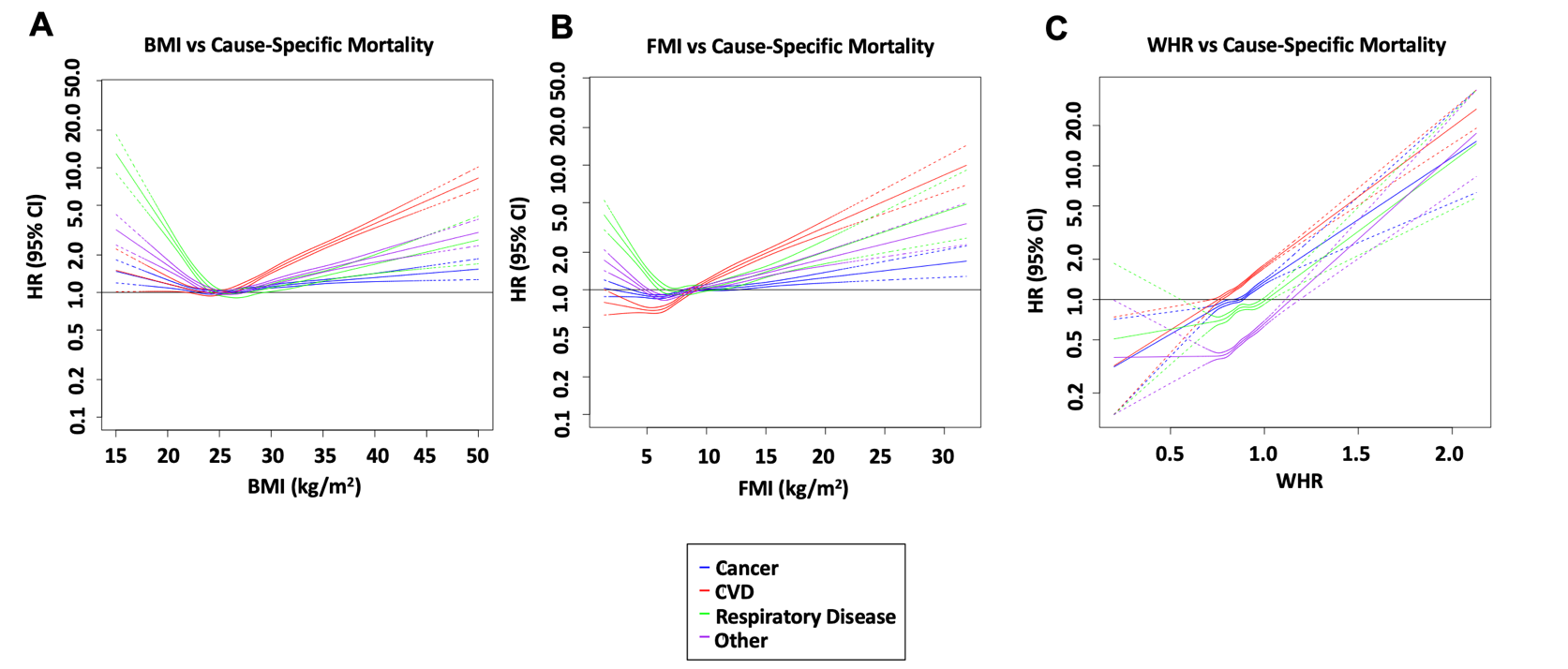

**Supplementary S11**: The relationship between a) BMI, b) FMI, and c) WHR respectively, and cause-specific mortality outcomes. BMI = body mass index, FMI = fat mass index, WHR = Waist-to-hip ratio, CVD = cardiovascular disease, HR = hazard ratio for all-cause mortality. Statistical significance for non-linearity was at a *p* < 0.05. N = 387,672. Against respiratory disease, the nadir for BMI and FMI were 26.0 and 7.43 kg/m^2^ respectively. Against other disease, the nadir for BMI and FMI were 25.5 and 6.55 kg/m^2^ respectively.

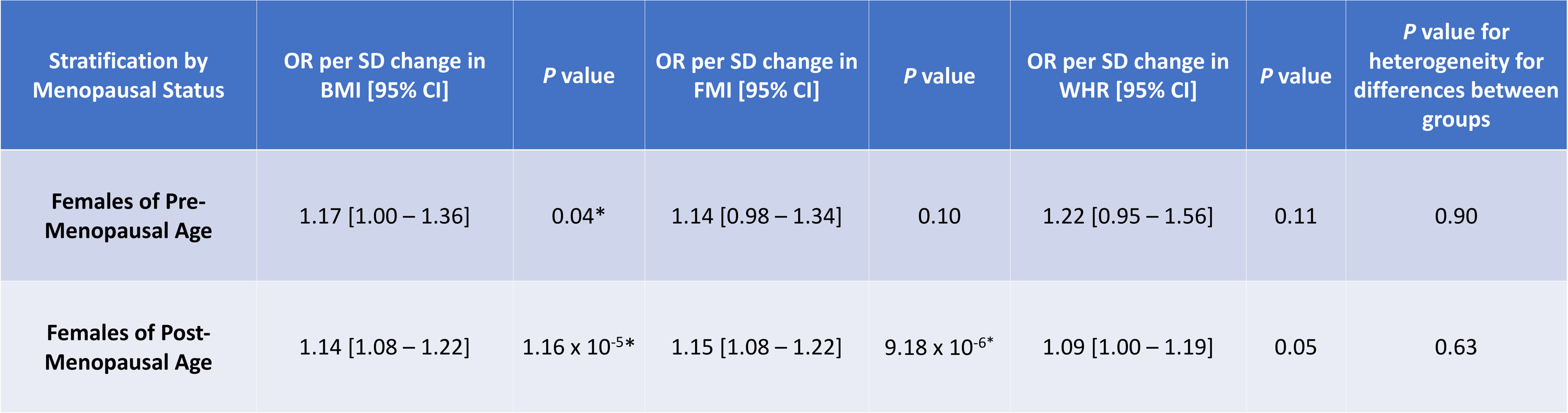

**Supplementary S12:** Linear mendelian randomization analyses comparing the effect of individual genetically-determined adiposity measures on all-cause mortality between pre- versus post-menopausal aged females in the UKB. Pre-menopausal age was defined as females aged 52 and younger, while post-menopausal age was defined as females aged 53 and older. All PRS were standardized for their effects on their corresponding traits (i.e. the BMI PRS was adjusted for its effect on BMI). Odds ratios (OR) indicate the effect of a 1 SD unit increase in adiposity measure on risk of all-cause mortality. Significance is considered at *p* < 0.05. BMI = body mass index, FMI = fat mass index, WHR = waist-to-hip ratio, OR = odds ratio, PRS = polygenic risk score, *P*_het_ = *p* value for heterogeneity from the fixed-effects general heterogeneity test. N_pre-menopausal_ = 2,300; N_post-menopausal_ = 18,263. The adjusted model was used. Asterisks (*) represent statistical significance.

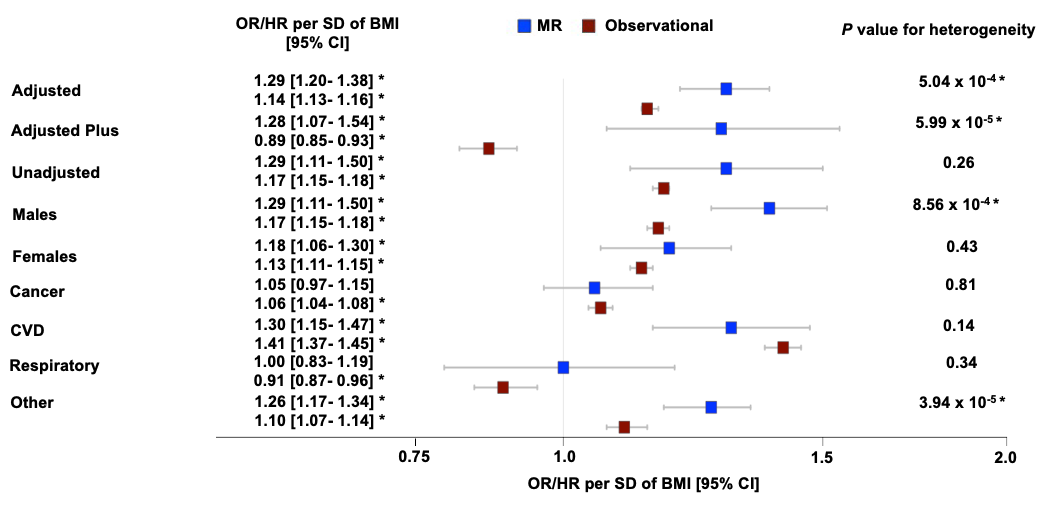

**Supplementary S13:** Comparison between epidemiologically-derived and MR-derived estimates for the BMI – all-cause mortality relationship. All PRS were standardized for their effects on their corresponding traits (e.g. the BMI PRS was adjusted for its effect on BMI). Hazard ratios (HR) indicate the effect of a 1 SD unit increase in BMI on risk of mortality. Odds ratios (OR) indicate the effect of a 1 SD unit increase in genetically-predicted BMI measure on risk of mortality. The adjusted model was used for all sex-specific and cause-specific mortality analyses. Significance is considered at *p* < 0.05. MR = mendelian randomization, BMI = body mass index, FMI = fat mass index, WHR = waist-to-hip ratio, CVD = cardiovascular disease, OR = odds ratio, HR = hazard ratio, PRS = polygenic risk score. N= 387,672 (adjusted, adjusted plus, unadjusted, and cause-specific mortality models with males and females combined, epidemiological analyses), N=50,594 (adjusted, adjusted plus, unadjusted, and cause-specific mortality models with males and females combined, MR analyses), N=177,340 (males only cohort, epidemiological analyses), N=210,332 (females only cohort, epidemiological analyses), N=30,031 (males only cohort, MR analyses), and N=20,563 (females only cohort, MR analyses). The *p* value for heterogeneity was obtained from the fixed-effects general heterogeneity test. Asterisks (*) represent statistical significance.

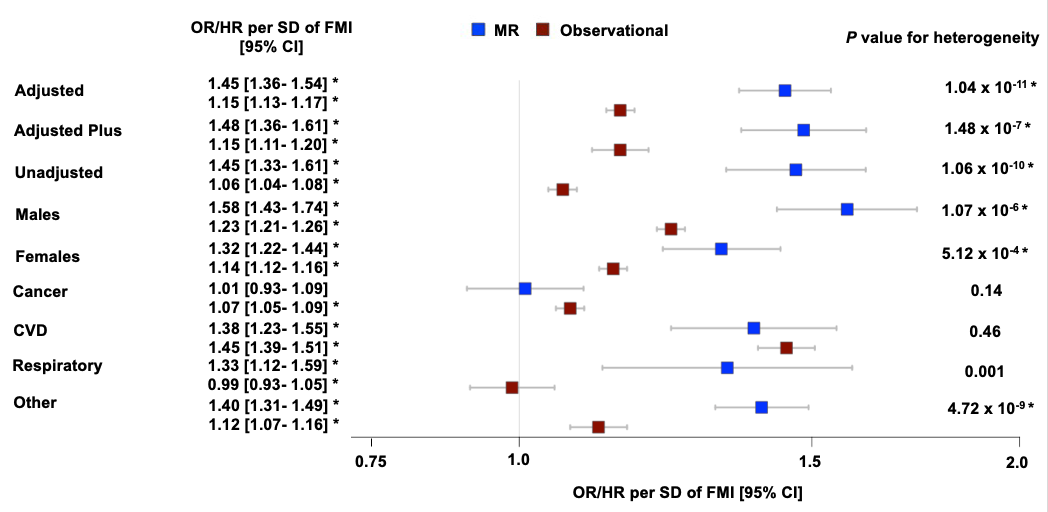

**Supplementary S14:** Comparison between epidemiologically-derived and MR-derived estimates for the FMI – all-cause mortality relationship. All PRS were standardized for their effects on their corresponding traits (e.g. the FMI PRS was adjusted for its effect on FMI). Hazard ratios (HR) indicate the effect of a 1 SD unit increase in FMI on risk of mortality. Odds ratios (OR) indicate the effect of a 1 SD unit increase in genetically-predicted FMI measure on risk of mortality. The adjusted model was used for all sex-specific and cause-specific mortality analyses. Significance is considered at *p* < 0.05. MR = mendelian randomization, BMI = body mass index, FMI = fat mass index, WHR = waist-to-hip ratio, CVD = cardiovascular disease, OR = odds ratio, HR = hazard ratio, PRS = polygenic risk score. N= 387,672 (adjusted, adjusted plus, unadjusted, and cause-specific mortality models with males and females combined, epidemiological analyses), N=50,594 (adjusted, adjusted plus, unadjusted, and cause-specific mortality models with males and females combined, MR analyses), N=177,340 (males only cohort, epidemiological analyses), N=210,332 (females only cohort, epidemiological analyses), N=30,031 (males only cohort, MR analyses), and N=20,563 (females only cohort, MR analyses). The *p* value for heterogeneity was obtained from the fixed-effects general heterogeneity test. Asterisks (*) represent statistical significance.

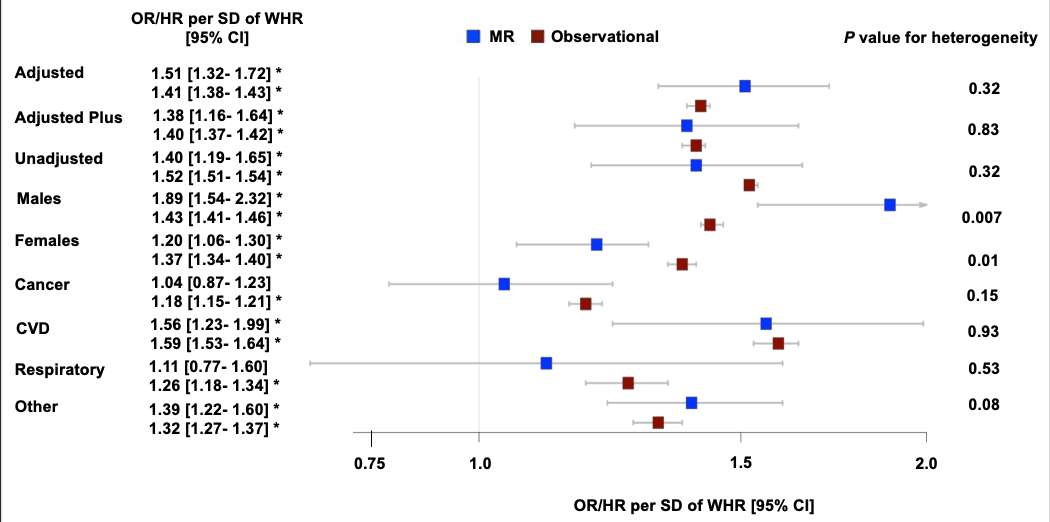

**Supplementary S15:** Comparison between epidemiologically-derived and MR-derived estimates for the WHR – all-cause mortality relationship. All PRS were standardized for their effects on their corresponding traits (e.g. the WHR PRS was adjusted for its effect on WHR). Hazard ratios (HR) indicate the effect of a 1 SD unit increase in WHR on risk of mortality. Odds ratios (OR) indicate the effect of a 1 SD unit increase in genetically-predicted WHR measure on risk of mortality. The adjusted model was used for all sex-specific and cause-specific mortality analyses. Significance is considered at *p* < 0.05. MR = mendelian randomization, BMI = body mass index, FMI = fat mass index, WHR = waist-to-hip ratio, CVD = cardiovascular disease, OR = odds ratio, HR = hazard ratio, PRS = polygenic risk score. N= 387,672 (adjusted, adjusted plus, unadjusted, and cause-specific mortality models with males and females combined, epidemiological analyses), N=50,594 (adjusted, adjusted plus, unadjusted, and cause-specific mortality models with males and females combined, MR analyses), N=177,340 (males only cohort, epidemiological analyses), N=210,332 (females only cohort, epidemiological analyses), N=30,031 (males only cohort, MR analyses), and N=20,563 (females only cohort, MR analyses). The *p* value for heterogeneity was obtained from the fixed-effects general heterogeneity test. Asterisks (*) represent statistical significance.
